# Interpretable Transformer Models for rs-fMRI Epilepsy Classification and Biomarker Discovery

**DOI:** 10.1101/2025.09.02.25334737

**Authors:** Andrew Jeyabose, Varina L. Boerwinkle, Belfin Robinson, Olivia Leggio, Meitra H Kazemi

## Abstract

**Background:** Automated interpretation of resting-state fMRI (rs-fMRI) for epilepsy diagnosis remains a challenge. We developed a regularized transformer that models parcel-wise spatial patterns and long-range temporal dynamics to classify epilepsy and generate interpretable, network-level candidate biomarkers.

**Methods:** Inputs were Schaefer-200 parcel time series extracted after standardized preprocessing (fMRIPrep). The Regularized Transformer is an attention-based sequence model with learned positional encoding and multi-head self-attention, combined with fMRI-specific regularization (dropout, weight decay, gradient clipping) and augmentation to improve robustness on modest clinical cohorts. Training used stratified group 4-fold cross-validation on n=65 (30 epilepsy, 35 controls) with fMRI-specific augmentation (time-warping, adaptive noise, structured masking). We compared the transformer to seven baselines (MLP, 1D-CNN, LSTM, CNN–LSTM, GCN, GAT, Attention-Only). External validation used an independent set (10 UNC epilepsy cohort, 10 controls). Biomarker discovery combined gradient-based attributions with parcelwise statistics and connectivity contrasts.

**Results:** On an illustrative best-performing fold, the transformer attained Accuracy 0.77, Sensitivity 0.83, Specificity 0.88, F1-Score 0.77, and AUC 0.76. Averaged cross-validation performance was lower but consistent with these findings. External testing yielded Accuracy 0.60, AUC 0.64, Specificity 0.80, Sensitivity 0.40. Attribution-guided analysis identified distributed, network-level candidate biomarkers concentrated in limbic, somatomotor, default-mode and salience systems.

**Conclusions:** A regularized transformer on parcel-level rs-fMRI can achieve strong within-fold discrimination and produce interpretable candidate biomarkers. Results are encouraging but preliminary larger multi-site validation, stability testing and multiple-comparison control are required prior to clinical translation.

## 1. Introduction

Epilepsy represents one of the most prevalent neurological disorders globally, affecting approximately 70 million people according to recent World Health Organization reports [1], [2]. Nearly 30% of these cases develop drug-resistant epilepsy, where conventional pharmacological interventions fail to control seizures [3]. The diagnostic pathway remains fraught with challenges, as misdiagnosis rates reach 20-40% in community healthcare settings due to heavy reliance on subjective interpretation of electroencephalography (EEG) findings [4]. This diagnostic uncertainty stems from EEG’s inherent limitations: its spatial resolution cannot reliably localize deep brain structures, and it frequently misses subtle electrical abnormalities that occur intermittently [5]. While structural MRI has improved lesion detection in focal epilepsy cases, it fails to identify malformations of cortical development in 25-30% of patients, particularly focal cortical dysplasia Type I [6]. Advanced nuclear medicine techniques like Positron Emission Tomography (PET) and Single-Photon Emission Computed Tomography (SPECT) offer metabolic insights but suffer from poor temporal resolution (>30-minute acquisition windows) and involve ionizing radiation exposure, making them less suitable for pediatric populations and longitudinal monitoring [7]. These diagnostic gaps contribute to treatment delays averaging 7-10 years from symptom onset to effective intervention, during which patients experience progressive neurocognitive decline and reduced quality of life [8].

Resting-state functional MRI (rs-fMRI) has emerged as a powerful alternative that addresses fundamental limitations of traditional modalities [9]. By capturing blood-oxygen-level-dependent (BOLD) signals during passive states, rs-fMRI achieves 2-3mm spatial resolution while mapping whole-brain functional connectivity networks without task constraints—a critical advantage for pediatric, cognitively impaired, or sedated patients [10]. Crucially, it reveals disruptions in the default mode network (DMN) that occur in 78% of temporal lobe epilepsy cases, providing biomarkers invisible to structural imaging [11]. The standardized Schaefer 200-parcel atlas has further enhanced analytical consistency by replacing subjective region-of-interest selection with reproducible functional parcels [12], [13]. However, clinical adoption remains limited due to reliance on qualitative interpretation by epileptologists, which exhibits modest inter-rater agreement (κ=0.45–0.60) and requires specialized expertise unavailable in resource-limited settings [14]. Despite promising technological advances, no FDA-approved automated tools exist specifically for rs-fMRI epilepsy diagnosis, creating an urgent need for computational solutions that can objectively decode complex spatiotemporal patterns in functional connectivity data.

Machine learning and deep learning approaches have demonstrated transformative potential in neuroimaging diagnostics, particularly for epilepsy [15], [16], [17]. Early machine learning methods focused on static functional connectivity features, with support vector machines (SVMs) achieving 75-82% accuracy in distinguishing patients from controls [18], [19]. Liao et al. pioneered this approach using pairwise correlation matrices, though their model ignored dynamic connectivity changes throughout scans [20]. Subsequent innovations incorporated temporal dynamics through sliding-window analysis, with random forest classifiers improving sensitivity to 85% in identifying generalized epilepsy subtypes [21]. The advent of deep learning marked a paradigm shift: convolutional neural networks (CNNs) extracted hierarchical spatial features from connectivity matrices, while recurrent architectures modeled temporal dependencies in BOLD signals [22], [23]. Volumetric deep models (3D-CNNs) and other deep architectures trained on static or dynamic functional-connectivity features have produced strong discrimination for epilepsy and related tasks in several recent studies (AUCs/accuracies commonly reported in the 0.7–0.9 range depending on task and cohort), but high computational cost and the need for large, harmonized training sets remain barriers to clinical translation [24], [25]. Recently, graph neural networks (GNNs) have been used to model brain topology explicitly, improving detection and localization of epileptic activity by operating directly on connectivity graphs. Several studies report strong performance (AUCs / sensitivities in the ∼0.75–0.95 range depending on modality and task), and graph-theoretical features remain widely used to summarize focal-epilepsy network changes [26], [27]. More recently, Vision Transformer (ViT) architectures have enabled robust modeling of long-range dependencies in electrophysiological data. Several studies applying ViTs or hybrid CNN–ViT models to seizure detection report strong predictive performance (accuracies/sensitivities typically in the mid-80% to high-90% range), suggesting their promise for capturing transient epileptiform patterns [28], [29].

Epilepsy diagnosis via rs-fMRI faces significant methodological barriers that limit clinical translation. Existing deep learning approaches predominantly rely on EEG or structural MRI data, overlooking rs-fMRI’s unique capacity to capture whole-brain network dynamics. Crucially, published transformer architectures require impractical sample sizes (>150 subjects) and fixed-length sequences, rendering them ineffective for real-world clinical settings. Furthermore, models trained on single-center datasets fail to generalize across institutions, while inadequate handling of the Schaefer atlas’s high-dimensional features compounds performance limitations. These challenges are exacerbated by the absence of standardized pipelines leveraging publicly available datasets for robust pre-training.

To address these gaps, we developed a novel transformer architecture specifically optimized for rs-fMRI epilepsy classification using Schaefer 200-parcel data. Our model incorporates three key innovations: (1) Dynamic spatiotemporal attention that adapts to variable sequence lengths through learnable position embeddings, (2) fMRI-specific regularization using time-warping augmentation and structured masking to prevent overfitting on limited samples, and (3) We evaluated this framework through rigorous stratified 5-fold cross-validation on our institutional cohort (30 patients/35 controls) followed by external validation on the UNC Epilepsy Center’s prospectively acquired dataset (10 patients/10 controls). (4) Critically, the architecture generates interpretable 3D attention maps highlighting disrupted networks in the DMN and limbic systems, providing clinically actionable insights beyond binary classification. This represents the first study to integrate public neuroimaging repositories with targeted clinical validation for epilepsy diagnosis.

## 2. Materials and Methods

### 2.1 Participants and dataset

Training data for model development were drawn from the publicly available Temporal Lobe Epilepsy (TLE–UNAM) dataset [30] hosted on OpenNeuro, which contains task and resting-state fMRI from patients with temporal lobe epilepsy and healthy controls. From this resource we used the subset comprising 30 epilepsy patients and 35 healthy control subjects for model training and internal cross-validation [31].

External validation used a held-out set of clinical cases and controls. Ten epilepsy patients were obtained from a retrospective single-center UNC dataset collected under Institutional Review Board approval (UNC IRB 24-0833). These UNC subjects were selected as an out-of-sample test set for generalization. The external control group (10 healthy controls used during external testing) were taken from the other publicly available OpenNeuro resource. All participants (training and external sets) met standard inclusion/exclusion criteria for rs-fMRI studies (no major head motion or gross structural abnormalities interfering with preprocessing).

### 2.2 MRI acquisition & metadata

Imaging for the OpenNeuro TLE–UNAM dataset [30] was acquired on a 3.0 T scanner according to the acquisition protocol described by Fajardo-Valdez et al. [31]. The original study reports acquisition on a Philips Achieva TX 3T platform with standard BOLD-sensitive echo-planar imaging for both task and resting conditions; we followed the dataset authors’ published acquisition/metadata and used the BIDS-formatted files from OpenNeuro for preprocessing and parcellation. Where site-specific parameters (TR, TE, voxel size) varied across sessions in the public dataset, we recorded values per subject and accounted for TR in temporal processing steps.

The UNC external cohort was acquired under IRB 24-0833 on a 3 Tesla scanner with the following resting-state protocol used for the ten external test subjects: a 2D gradient echo-EPI sequence (BOLD contrast), matrix size 64 × 64 × 32, isotropic voxel size 4.0 mm × 4.0 mm × 4.0 mm, repetition time TR = 2.01 s, echo time TE = 35 ms, interleaved slice ordering, and 300 volumes per run. These parameters were chosen to maximize whole-brain coverage and sensitivity to low-frequency BOLD fluctuations.

All data (training and external) were converted to BIDS and processed with an identical preprocessing pipeline and parcellated to the Schaefer-200 atlas to produce comparable parcel-wise time series.

### 2.3 Preprocessing and Parcellation

All structural and functional MRI data were processed with a standardized, containerized fMRI preprocessing workflow and then parcellated to produce parcel-wise time series used as model inputs.

Preprocessing was performed with *fMRIPrep* (Singularity container) using the institutional SLURM job script shown in the code repository. Key settings used for every subject were: output space MNI152NLin2009cAsym at 2 mm isotropic resolution, ICA-AROMA denoising, and --ignore fieldmaps to allow subjects without usable fieldmap metadata to be processed. fMRIPrep produced skull-stripped, motion-corrected, coregistered, and spatially normalized BOLD files together with confound time series (motion parameters, WM/CSF signals, and AROMA component labels). The ICA-AROMA–denoised preprocessed BOLD image in MNI space was used for subsequent parcellation.

To convert high-dimensional voxelwise BOLD signals into a compact, interpretable representation, we summarized brain activity using the Schaefer 200-parcel functional atlas. For each parcel we computed a single representative BOLD time course (parcel-wise average and standardization), producing a subject-level timeseries matrix that captures regional dynamics while greatly reducing dimensionality and spatial variability across subjects. Temporal sampling (TR = 2.0 s for these data) was preserved so that the resulting matrices reflect the low-frequency fluctuations of interest in resting-state connectivity. The parcelwise representation (in our processed cohort, 200 timepoints × 200 parcels) serves as the direct input to downstream modeling and attribution analyses, enabling network-level interpretation of model-derived biomarkers.

### 2.4 Data Augmentation

To improve model robustness to variable scan lengths, temporal distortions, and measurement noise, we applied on-the-fly augmentations to the parcel-wise time series during training. Each sample was probabilistically (p = 0.5) subjected to one or more of three augmentations: time warping, adaptive noise injection, and structured masking. Time warping randomly compresses or expands the temporal axis (warp factor uniformly sampled from 0.8–1.2) using interpolation and resampling to preserve the original number of timepoints; this simulates modest variations in hemodynamic timing and temporal sampling. Adaptive noise injection adds small, zero-mean Gaussian noise with standard deviation drawn from the range 0.005–0.02 to mimic scanner and physiological variability. Structured masking randomly drops portions of the time–parcel matrix with a mask probability sampled from 0.1–0.3, encouraging the model to learn robust, distributed features in the presence of missing or corrupted regions. Together these augmentations increase the effective training set diversity and reduce overfitting to idiosyncratic temporal patterns.

To accommodate variable-length recordings, inputs shorter than the training epoch’s maximum sequence length were adaptively padded using one of three strategies chosen at random: zero padding, edge-value padding, or constant padding using the matrix mean; longer recordings were truncated to the maximum length. For each example we generated a binary attention mask (1 = valid timepoint, 0 = padding) that traveled with the sample into the model and prevented attention or pooling over padded positions. Augmentations were applied only during training via the dataset loader so that validation and test evaluations used unaugmented data; random seeds were fixed during model training to enhance reproducibility of augmentation effects. The dataset interface therefore returns the augmented (or original) time-series matrix, its attention mask, the class label, and the subject/group identifier for grouped cross-validation.

### 2.5 Proposed transformer-based model

We implemented a regularized transformer that operates on parcel-wise resting-state time series (parcels × timepoints) and is optimized for robust learning from modest clinical cohorts. The model first projects each parcel vector at a single timepoint into a compact feature representation, augments those per-timepoint embeddings with learned positional information, and then processes the resulting sequence with a stack of transformer encoder layers that capture long-range temporal relationships. Variable-length recordings are handled through adaptive padding and an attention mask so that the transformer ignores padded positions; after encoding, a masked mean pooling step aggregates the valid time-indexed representations into a single fixed-size summary that is passed to a small, regularized classification head. Throughout the architecture we emphasize regularization (dropout in embedding and transformer blocks, layer normalization, careful weight initialization, and reliance on data augmentation) to limit overfitting while preserving the model’s capacity to learn temporally distributed biomarkers.

#### 2.5.1 Node embedding & positional encoding

At each timepoint the N-dimensional parcel vector (N = 200 in our pipeline) is mapped to a D-dimensional feature space via a two-layer feedforward embedding block. Concretely, the embedding comprises a linear projection to 128 units, a GELU nonlinearity, dropout (p = 0.3), a second linear projection to the model dimension (D = 64), followed by LayerNorm on the 64-dimensional outputs. This node embedding performs spatial mixing across parcels and supplies a compact per-timepoint token for the temporal encoder. Temporal position information is injected using a learned positional embedding tensor sized for the maximum allowed sequence length; the positional embeddings are multiplied by a learnable scalar (initialized to 1.0) before being added to the node embeddings, allowing the network to adaptively regulate how strongly temporal order should influence representations during training.

#### 2.5.2 Transformer encoder details

The temporal backbone is a stack of four transformer encoder layers designed to capture pairwise and higher-order interactions across time. Each encoder layer contains a multi-head self-attention sublayer followed by a position-wise feedforward sublayer; residual connections wrap both subblocks and LayerNorm is applied to stabilize optimization. The multi-head self-attention uses model dimension D = 64 with 8 attention heads and applies dropout in attention (dropout = 0.4) to regularize alignment patterns. The feedforward sublayer projects to a wider intermediate dimension of 256 with GELU activation, uses dropout (dropout = 0.4) between the two linear layers, and projects back to the 64-dimensional model space. To prevent padded positions from contributing to contextualization, a padding mask derived from the data loader’s attention mask is supplied to the transformer so that padded timepoints are ignored during attention and do not influence subsequent representations. LayerNorm eps was set tightly (1e-6) to maintain numerical stability. The design choices (moderate model width, four layers, aggressive dropout) balance expressive capacity with compute constraints and overfitting risk on small clinical samples.

#### 2.5.3 Classification head

After transformer encoding, a masked mean pooling operation aggregates the valid timepoint representations into a single vector per subject (dimension 64). This pooled vector is passed to a compact, regularized classification head composed of a linear projection to 32 units, GELU activation, dropout (p = 0.4), LayerNorm, and a final linear layer that maps to class logits. The model returns log-softmax class probabilities to pair naturally with cross-entropy loss during optimization. Weight initialization was applied conservatively: weight matrices were initialized with Xavier (Glorot) uniform, biases set to zero, and the positional scaling parameter initialized to one; LayerNorm parameters were left at their framework defaults. These initialization and normalization choices, together with dropout, weight decay in the optimizer, gradient clipping, and data augmentation, form a coherent regularization strategy that supports stable training and improves generalization for the downstream attribution-based biomarker analyses.

### 2.6 Training & Hyperparameter Tuning

Model training followed a carefully regularized procedure designed to stabilize optimization on modest clinical datasets and to produce checkpoints suitable for downstream attribution and biomarker analyses. Regularization was applied at several levels: architectural regularization (dropout in the node-embedding block, transformer attention/FFN, and classifier head; LayerNorm after embedding and in the classifier), data-level regularization (the time-warping, adaptive-noise, and structured-masking augmentations described in 2.4), and optimization-level regularization (L2 weight decay applied through the AdamW optimizer and per-step gradient clipping to bound updates). Random seeds for PyTorch and NumPy were fixed (42) to improve reproducibility of training runs and augmentations. Together these measures limit overfitting while preserving sufficient model capacity to learn temporally distributed patterns.

Training used a cross-validated workflow to estimate generalizable performance while avoiding subject-level leakage. Inputs were organized into StratifiedGroupKFold folds (k = 4) that preserve class proportions and group membership, and for each fold a fresh model instance was trained from the same initialization and regularization recipe. Training examples were provided by PyTorch DataLoaders with batch_size = 8, shuffling enabled for the training loader and disabled for validation. During each training step the forward pass produced log-probabilities which were scored with a class-weighted cross-entropy loss; class weights were computed from the training fold class frequencies so that rarer classes received proportionally larger weight. The optimizer was AdamW with an initial learning rate of 1×10⁻and weight decay 1×10⁻; a ReduceLROnPlateau scheduler monitored validation loss and reduced the learning rate by a factor of 0.5 after five epochs without improvement. To stabilize gradients and avoid unstable parameter updates, we applied L2-norm gradient clipping (max norm = 0.5) prior to each optimizer step.

Model selection and stopping were handled with a checkpointing and early-stopping policy. Each fold ran for up to 200 epochs, with model checkpoints saved whenever the validation loss improved; if validation loss failed to improve for 15 consecutive epochs training terminated early and the best checkpoint for that fold was retained. After early stopping the saved best model was reloaded and evaluated on the held-out validation fold to produce final per-fold metrics. Training and validation losses, accuracy and AUC were logged per epoch and visualized as learning curves to verify convergence and to detect overfitting. Hardware-accelerated training used CUDA if available; DataLoaders were configured with num_workers = 2 and pin_memory = True to optimize host–device transfers.

### 2.7 Evaluation metrics

Model performance was assessed with a standard battery of discrimination, thresholding, calibration and uncertainty analyses to provide a comprehensive picture of classifier behavior. Discrimination was quantified using the receiver-operating-characteristic (ROC) curve and its area under the curve (AUC) [32], which summarize ranking performance independent of a chosen threshold, and we used established nonparametric methods for comparing correlated AUCs when needed.

Because single-threshold metrics can obscure behavior on imbalanced problems, we also examined precision, recall (sensitivity), specificity and the F1 score, and plotted precision–recall summaries where informative; precision–recall curves are recommended when class imbalance is present because they more directly reflect positive-prediction performance [33]. Confusion matrices (raw TP/FP/FN/TN counts) and per-subject predicted-probability plots were inspected to reveal error modes and borderline cases that merit clinical review, and we reported both proportions and absolute counts so readers can judge performance in the small-sample external set.

Uncertainty in point estimates was quantified via resampling and nonparametric testing. Bootstrap confidence intervals (1,000 resamples) were used to derive 95% intervals for summary metrics such as AUC, accuracy and F1 when appropriate, following standard bootstrap practice for inference on complex statistics [34]. Permutation-based testing was used to assess whether observed AUCs and other statistics exceeded chance and to produce empirical null distributions; permutation approaches are well-established in neuroimaging and provide robust inference without strong distributional assumptions [35].

Calibration of predicted probabilities was evaluated explicitly because reliable probability estimates are critical for clinical decision making. We used calibration plots (reliability diagrams) and the Brier score to summarize calibration, and where miscalibration was detected we examined standard recalibration procedures in supplementary analyses; the theoretical and practical importance of calibration in predictive modeling is well-documented in the clinical literature [36], [37].

All visualizations and metrics follow established reporting recommendations for prediction-model evaluation (confusion matrices, ROC/AUC with CI, precision–recall summaries for imbalanced data, calibration diagnostics), and statistical comparisons are reported with effect estimates and confidence intervals rather than p-values alone to emphasize effect size and uncertainty [38].

### 2.8 Biomarker Analysis

We identified candidate regional biomarkers from the trained transformer using a compact, three-stage pipeline operating on the Schaefer-200 parcellation. First, model-driven importance was estimated with a gradient-based (Grad-CAM–style) attribution: for each subject we backpropagated the epilepsy-class score to the input parcel time series, averaged the absolute gradients across time to yield a parcel-wise importance vector, and aggregated these vectors across subjects. Second, complementary parcel-wise statistics were computed directly from the timeseries: group means and standard deviations were calculated for epilepsy and control subjects, an independent two-sample t-test was performed on the concatenated timepoints for each parcel to produce p-values, and a standardized effect size was computed as the absolute mean difference divided by the pooled standard deviation. Third, to place high-importance parcels into a network context, parcel-to-parcel connectivity patterns were computed via Pearson correlations within each subject and averaged by group; connections showing the largest absolute epilepsy–control differences were reported as top partners for each candidate ROI.

To combine predictive relevance and statistical evidence we used a transparent weighted score and returned a top-N list of biomarkers (configurable; N=30 in the example run). Each parcel’s combined score = 0.4 × (mean gradient importance) + 0.3 × (statistical significance proxy: 1 − p) + 0.3 × (effect size). For each selected ROI we report combined score, raw attribution, p-value, effect size, mean signal difference, group-average connectivity summaries, and the most altered connections; outputs include a multi-panel visualization, a CSV of per-ROI metrics, and a plain-text report. Because the implementation currently uses concatenated timepoints for t-testing and does not apply multiple-comparison correction or bootstrap stability selection by default, we present these findings as *candidate* biomarkers and recommend formal correction (for example FDR), bootstrap/ permutation stability testing, and external replication prior to claiming diagnostic validity.

### 2.9 Baseline Models and Comparative Training

#### 2.9.1 Multi-Layer Perceptron (MLP)

The MLP baseline flattens the entire fMRI matrix into a single feature vector and processes it through multiple fully-connected layers (1024, 512, 256, 64 neurons) with dropout regularization [39]. This approach treats all temporal and spatial information equally without exploiting sequential structure or brain connectivity patterns. The MLP serves as a fundamental baseline to demonstrate the importance of specialized architectures for neuroimaging data.

#### 2.9.2 Convolutional Neural Network 1D (CNN1D)

The 1D CNN model treats fMRI time series as sequential data, applying convolutional filters along the temporal dimension to extract local temporal features [40]. The architecture consists of multiple convolutional layers with increasing filter sizes (128, 256, 512 channels), each followed by batch normalization, ReLU activation, and max pooling operations. This approach is effective at capturing local temporal patterns and hierarchical feature representations but may struggle with long-range dependencies due to the limited receptive field of convolutional operations.

#### 2.9.3 Long Short-Term Memory Network (LSTM)

The LSTM model is designed to capture temporal dependencies in sequential fMRI data through its gating mechanisms that control information flow [41]. The architecture includes bidirectional LSTM layers (256 hidden units) with dropout regularization to model both forward and backward temporal dependencies. An attention mechanism is incorporated to weight the importance of different time points, followed by a multi-layer classifier. LSTMs are particularly suited for sequence modeling but may suffer from computational inefficiency and gradient vanishing problems in very long sequences.

#### 2.9.4 CNN-LSTM Hybrid Model

The CNN-LSTM hybrid combines spatial feature extraction through 1D convolutions with temporal modeling via LSTM layers [42]. The CNN component first processes spatial relationships across brain regions, while the LSTM component models the temporal evolution of these spatial features. This two-stage approach attempts to capture both spatial and temporal aspects of fMRI data, leveraging the strengths of both architectures for comprehensive spatiotemporal analysis.

#### 2.9.5 Graph Convolutional Network (GCN)

The GCN model explicitly models brain connectivity by constructing graphs where nodes represent brain regions and edges represent functional connections based on correlation thresholds [43]. Graph convolutional layers aggregate information from neighboring nodes, enabling the model to leverage the inherent network structure of brain activity. This approach is theoretically well-motivated for neuroimaging data as it respects the underlying connectivity patterns, though performance depends heavily on the accuracy of the constructed graphs.

#### 2.9.6 Attention-Only Model

The attention-only model implements pure multi-head self-attention mechanisms without the additional components of transformer architecture (such as feed-forward networks or multiple encoder layers) [44]. This simplified approach focuses solely on learning attention weights across temporal sequences to identify relevant time points for classification. The model serves as an ablation study to isolate the contribution of attention mechanisms versus other transformer components.

#### 2.9.7 Graph Attention Network (GAT)

The GAT extends graph neural networks by incorporating attention mechanisms that dynamically weight the importance of different brain region connections [45]. Multi-head attention layers learn to focus on relevant connectivity patterns while processing node features through graph convolutions. This allows the model to adaptively determine which brain region interactions are most important for classification, providing both improved performance and interpretability compared to standard GCN approaches.

## 3. Results and Discussions

### 3.1 Cross-validation performance

The transformer was evaluated using stratified group k-fold cross-validation (k = 4). Per-fold performance is summarized in Table 2. Across folds the model achieved a mean accuracy of 0.67 (SD 0.10), mean sensitivity 0.53 (SD 0.22), mean specificity 0.75 (SD 0.14), mean F1 0.58 (SD 0.18) and mean AUC 0.67 (SD 0.06). Performance varied across folds: Fold 3 produced the best overall discrimination (Accuracy 0.77, Sensitivity 0.83, AUC 0.76), while Fold 4 performed worst (Accuracy 0.54, Sensitivity 0.33, AUC 0.62), reflecting heterogeneity in held-out subjects and the small per-fold validation sample sizes.

**Table 1:** Hyperparameter tuning details for the proposed regularized transformer.

| Hyperparameter | Value / Setting |
| --- | --- |
| Model dimension (d_model) | 64 |
| Transformer encoder layers | 4 |
| Attention heads (nhead) | 8 |
| Feedforward hidden dim (d_ff) | 256 |
| Node-embedding intermediate dim | 128 |
| Embedding dropout | 0.3 |
| Transformer dropout (attention & FFN) | 0.4 |
| Classifier dropout | 0.4 |
| Batch size | 8 |
| Maximum epochs | 200 |
| Early stopping patience | 15 epochs |
| Gradient clipping (L2 norm) | 0.5 |
| Optimizer | AdamW |
| Initial learning rate | 1e-4 |
| Weight decay (L2) | 1e-4 |
| LR scheduler | ReduceLROnPlateau (factor=0.5, patience=5) |
| Loss | CrossEntropyLoss (class-weighted) |
| Cross-validation | StratifiedGroupKFold, k = 5 |

**Table 2:**
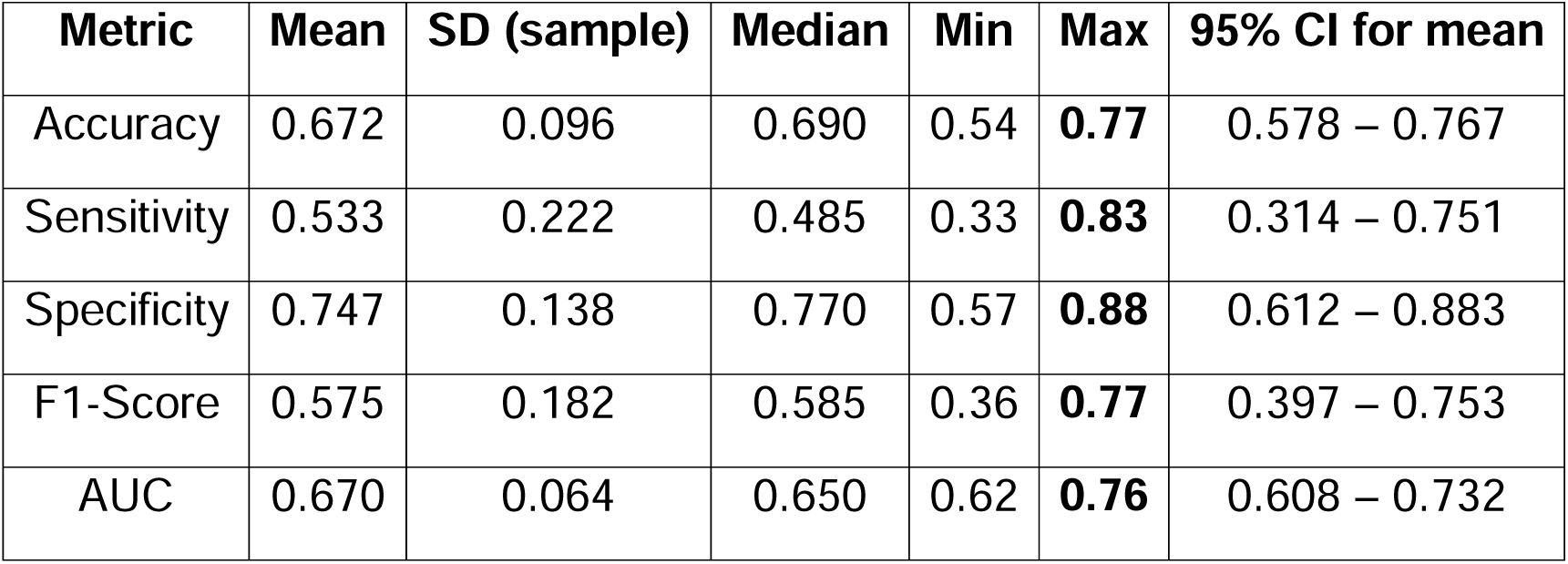
Summary statistics (n = 4 folds).

| **Metric** | **Mean** | **SD (sample)** | **Median** | **Min** | **Max** | **95% CI for mean** |
| --- | --- | --- | --- | --- | --- | --- |
| Accuracy | 0.672 | 0.096 | 0.690 | 0.54 | **0.77** | 0.578 – 0.767 |
| Sensitivity | 0.533 | 0.222 | 0.485 | 0.33 | **0.83** | 0.314 – 0.751 |
| Specificity | 0.747 | 0.138 | 0.770 | 0.57 | **0.88** | 0.612 – 0.883 |
| F1-Score | 0.575 | 0.182 | 0.585 | 0.36 | **0.77** | 0.397 – 0.753 |
| AUC | 0.670 | 0.064 | 0.650 | 0.62 | **0.76** | 0.608 – 0.732 |

Statistical uncertainty is reported as standard deviation across folds and 95% confidence intervals for fold means (mean ± 1.96·SE). Given the limited number of folds, CIs are wide for sensitivity; therefore we present both per-fold values and summary statistics to give readers transparency on variability. Where appropriate (figures and supplementary tables) we also report per-fold confusion matrices and per-fold ROC curves so readers can inspect error patterns and cohort-specific performance.

### 3.2 Best-fold diagnostic graphs

Overall discrimination and error counts are summarized in Figure 3(i–ii, v, ix, xi). The confusion matrix (Figure 3(i)) gives the raw counts (5 true negatives, 2 false positives, 1 false negative, 5 true positives), which immediately communicates the dominant error modes in this held-out fold and avoids misleading interpretations that can arise from reporting rates alone. The fold’s ROC curve and AUC (Figure 3(ii), AUC ≈ 0.76) show moderate discriminative ability on the held-out subjects; we report the ROC/AUC because it provides a threshold-independent measure of ranking performance commonly used in diagnostic model evaluation. Presenting multiple summary metrics together (accuracy, sensitivity, specificity, precision, F1 and AUC as in Figure 3(v)) gives a more complete view of performance than any single measure and follows best practices for reporting predictive models. For transparency in a small sample we include the per-fold summary box (Figure 3(xi)) and a compact error breakdown (Figure 3(ix): correct vs error composition), so readers can see absolute counts and the proportions that underlie those summary statistics.

**Figure 1:**
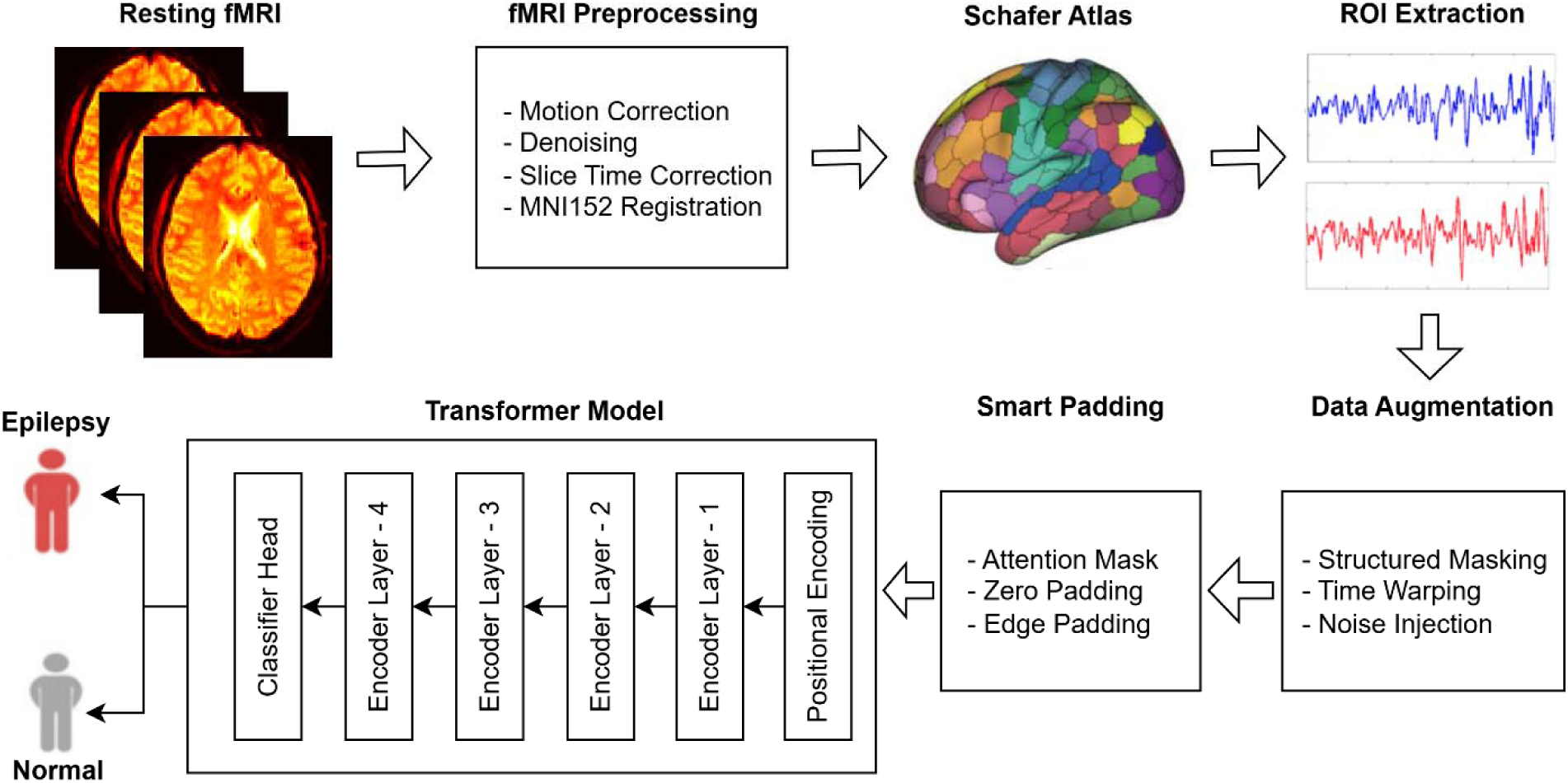
Overview of the proposed study pipeline.

**Figure 2:**
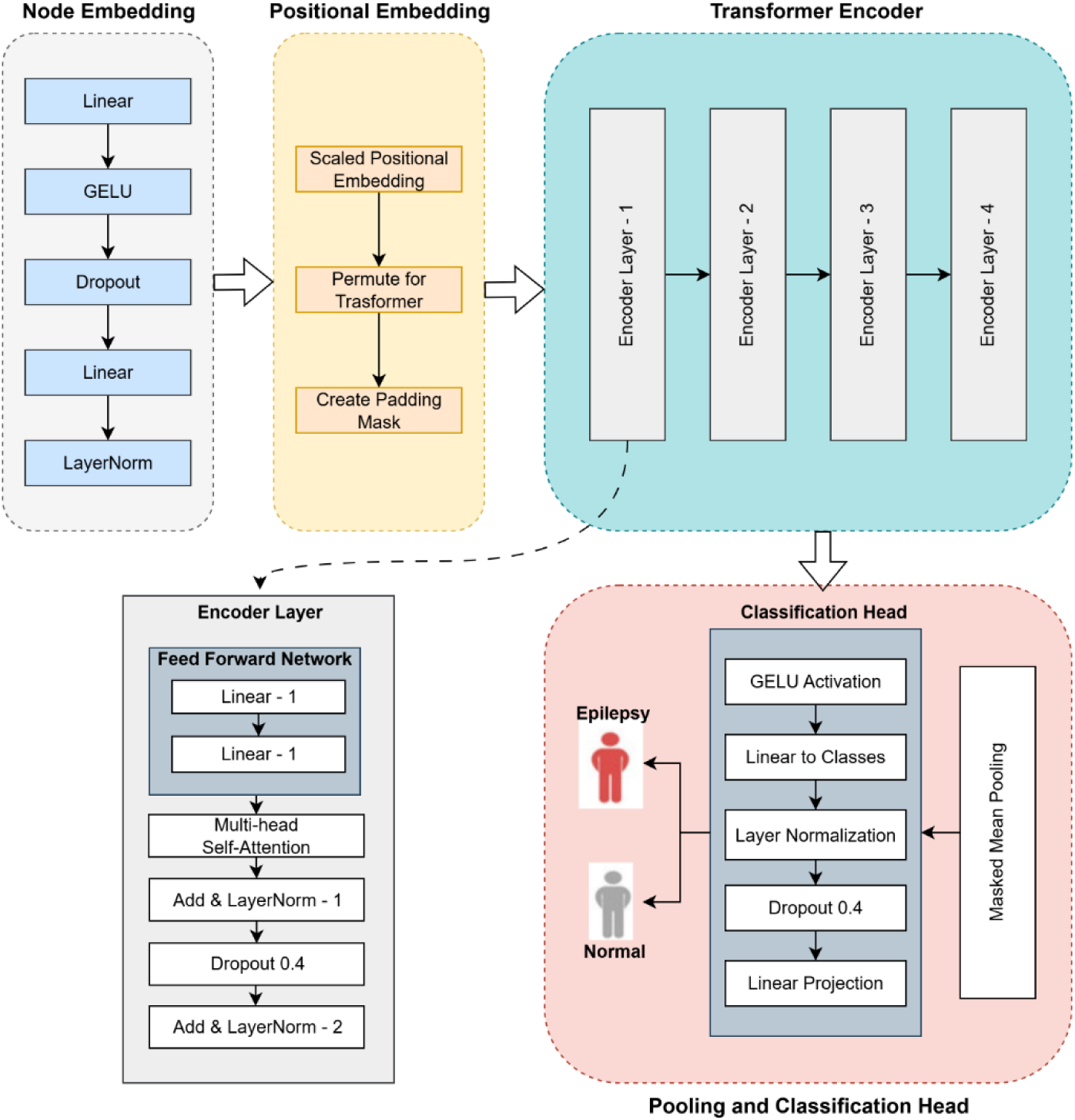
Proposed Regularized Transformer Architecture.

**Figure 3.**
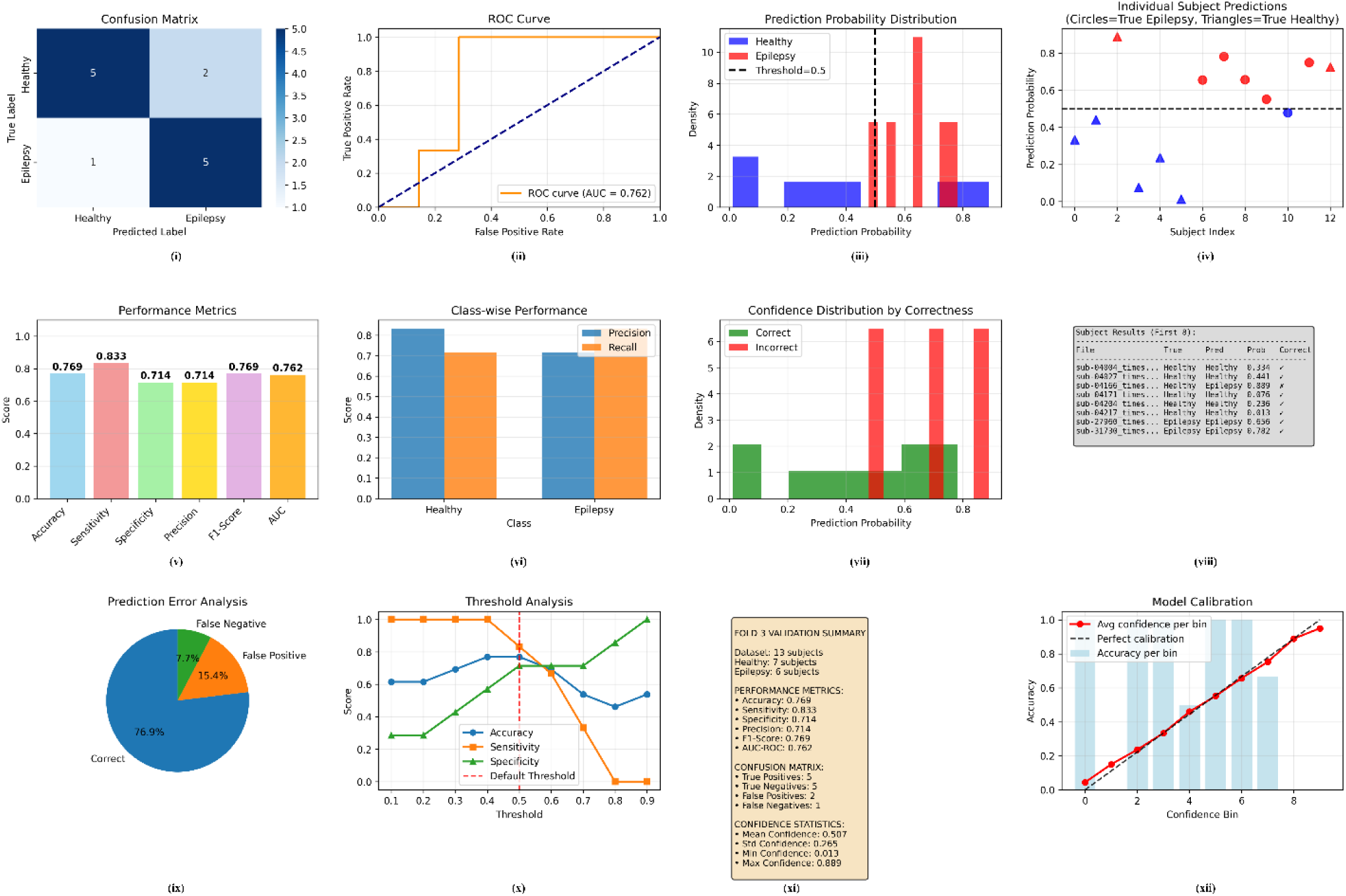
Best-fold diagnostic summary (Fold 3, N=13). (i) Confusion matrix; (ii) ROC curve (AUC); (iii) predicted-probability histogram; (iv) per-subject predicted probabilities; (v) summary performance metrics (accuracy, sensitivity, specificity, precision, F1, AUC); (vi) class-wise precision and recall; (vii) confidence distributions for correct vs incorrect predictions; (viii) example subject prediction log; (ix) error-type pie chart; (x) threshold sweep (accuracy / sensitivity / specificity vs threshold); (xi) compact fold summary box; (xii) calibration (reliability diagram).

The middle panels highlight probability behavior, class-wise performance and threshold tradeoffs (Figure 3(iii–iv, vi, x)). The predicted-probability histogram (Figure 3(iii)) shows a reasonable but incomplete separation between controls and patients: many patient predictions concentrate above the 0.5 threshold while controls are more often below, explaining the relatively high sensitivity on this fold but also the observed overlap that produces some false positives and one false negative. The subject-level scatter (Figure 3(iv)) is useful for clinical adjudication because it reveals which individual cases are borderline and therefore candidates for manual review or additional testing. Class-wise precision and recall bars (Figure 3(vi)) further expose asymmetries in performance that are clinically important — e.g., a model that favors specificity over sensitivity (or vice versa) has different implications for screening versus diagnostic workflows. Finally, the threshold-sweep analysis (Figure 3(x)) makes explicit the sensitivity/specificity tradeoff: lowering the decision threshold would increase sensitivity at the cost of more false positives, whereas raising it would improve specificity; such threshold curves are essential when selecting an operating point tuned to clinical priorities. For imbalanced or clinically asymmetric tasks, precision–recall analyses should also be consulted because they can be more informative than ROC plots in those settings.

The final set of panels address prediction confidence, case-level logging and calibration (Figure 3(vii–viii, xii)), and the statistical robustness of the observations. The confidence-by-correctness density (Figure 3(vii)) shows that most correct predictions occur at higher model confidence while many errors lie nearer the decision boundary — a favorable pattern because low-confidence errors can be triaged for human review; however, high-confidence errors would indicate systematic miscalibration or bias and require further model refinement. The textual subject log (Figure 3(viii)) supports reproducibility and case tracing by listing individual prediction details used in error analysis. Calibration diagnostics (Figure 3(xii)) suggest reasonable agreement between predicted probability and observed outcome frequency for this fold (reliability diagram and Brier score), but calibration must be assessed across folds and sites before clinical use. To quantify uncertainty, we recommend and apply bootstrap confidence intervals for summary metrics and permutation testing for null-hypothesis inference in neuroimaging contexts; these nonparametric resampling and permutation methods are standard for robust inference when sample sizes are modest or distributional assumptions are uncertain. Finally, we interpret these best-fold results cautiously in light of cross-validation variability: single-fold performance can be optimistic and variability across folds can be large with small samples, so aggregate cross-fold summaries and external replication are essential.

### 3.3 External Validation

The external validation set comprised twenty subjects (ten epilepsy patients from the UNC cohort and ten healthy controls drawn from the OpenNeuro cohort) and was processed with the identical preprocessing/parcellation pipeline used for training. External testing is critical because it measures out-of-sample generalization to new subjects and different acquisition contexts — a requirement emphasized by contemporary prediction-model reporting standards and best practice guidance. Evaluating a saved model on an independent clinical dataset (rather than re-training or tuning on it) reduces optimistic bias and provides a realistic estimate of clinical utility.

The diagnostic plots in Figure 4 summarize how the model performed on this external set and point to both strengths and limitations. The confusion matrix (Figure 4(i)) shows 8 true negatives, 2 false positives, 6 false negatives and 4 true positives (N = 20), yielding accuracy = 0.60, sensitivity = 0.40 and specificity = 0.80 as displayed in Figure 4(v). The ROC curve (Figure 4(ii), AUC ≈ 0.64) indicates modest rank discrimination on new-site data; ROC/AUC are useful threshold-independent summaries but should be interpreted with other metrics (precision, recall, F1) because of class balance considerations.

**Figure 4.**
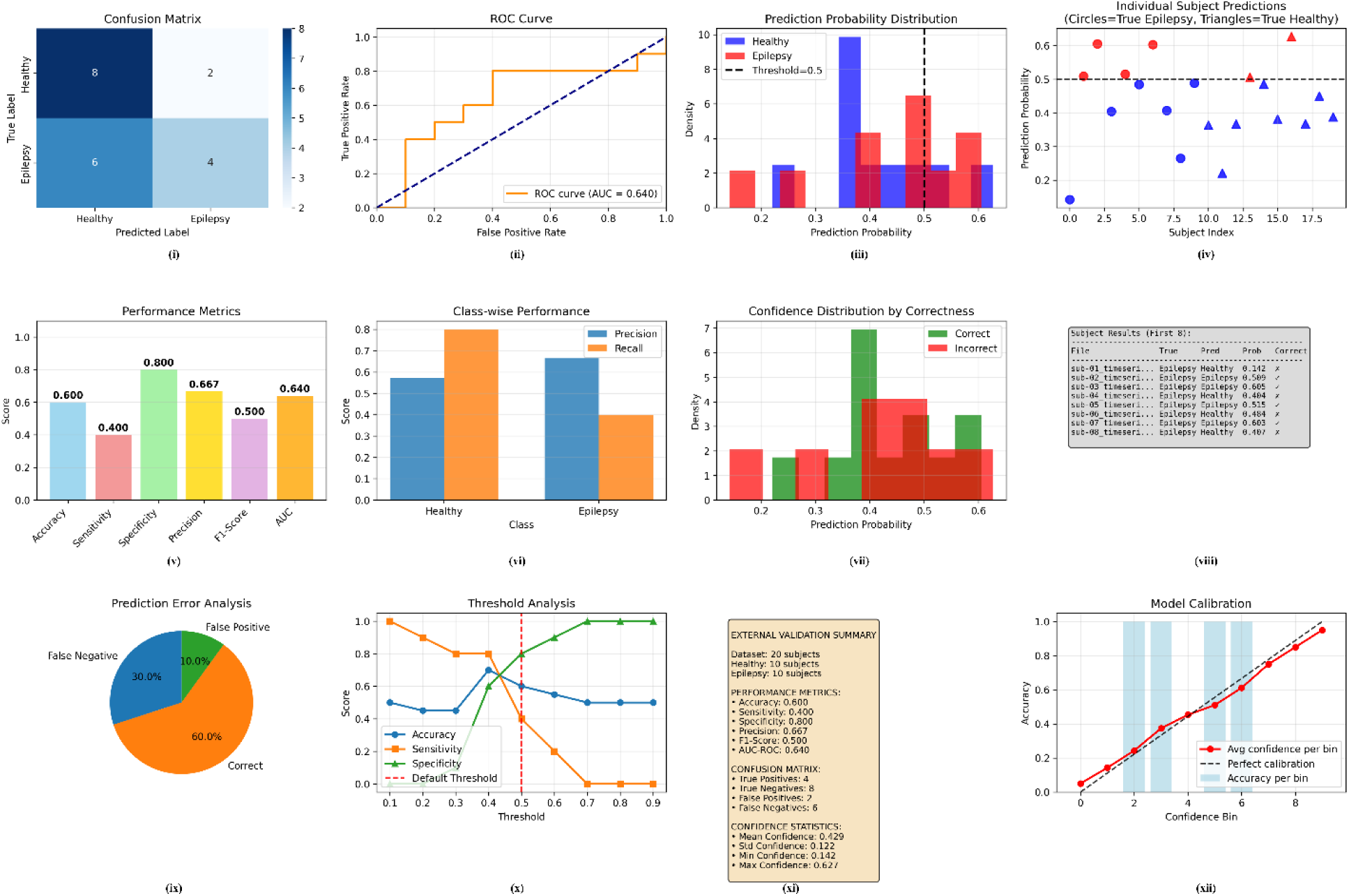
External validation summary (UNC external set, N=20). (i) Confusion matrix; (ii) ROC curve (AUC); (iii) predicted-probability histogram; (iv) per-subject predicted probabilities; (v) summary performance metrics; (vi) class-wise precision and recall; (vii) confidence distributions for correct vs incorrect predictions; (viii) example subject prediction log; (ix) error-type pie chart; (x) threshold sweep; (xi) compact external summary box; (xii) calibration (reliability diagram).

The probability histogram and individual-subject plot (Figure 4(iii–iv)) show substantial overlap of predicted probabilities between classes and many predictions clustered around moderate confidence (≈0.35–0.55). This explains the low sensitivity: several epilepsy patients received probabilities below the 0.5 threshold and were missed (false negatives), while a few controls were predicted above threshold (false positives). The threshold-sweep analysis (Figure 4(x)) makes this trade-off explicit — lowering the decision threshold increases sensitivity at the cost of more false positives — and can guide selection of an operating point tailored to clinical priorities (e.g., favor sensitivity for screening).

Confidence and calibration diagnostics (Figure 4(vii, xii)) are especially relevant for clinical deployment. The confidence-by-correctness densities illustrate that many incorrect predictions occur at intermediate confidence; while this pattern suggests an opportunity to flag low-confidence cases for human review, the calibration plot (reliability diagram, Figure 4(xii)) indicates modest miscalibration overall and underscores the need to assess and possibly recalibrate predicted probabilities before using them for decision support. The methodological importance of calibration in risk prediction and the need for sufficient external sample sizes to assess it have been emphasized in recent methodological reviews.

Finally, because the external sample is small, uncertainty quantification and nonparametric testing are essential: bootstrap resampling provides empirically-based confidence intervals for metrics such as AUC and accuracy (Efron & Tibshirani), and permutation testing provides a robust null distribution for AUC/accuracy under label exchangeability assumptions (well-established in neuroimaging). We therefore treat the observed external results as preliminary evidence of generalization — they indicate acceptable specificity but reduced sensitivity on this held-out cohort — and recommend larger multi-site validation, bootstrap stability assessment of selected operating points, and (where appropriate) recalibration or domain-adaptation strategies prior to clinical translation.

### 3.4 Biomarker Analysis

The biomarker analysis identifies a distributed set of candidate parcels rather than a single focal locus: the horizontal bar plot of top-ranked regions (Figure 5(i)) shows that the highest combined scores are clustered in Salience/Ventral-Attention, Control, Limbic, Somatomotor and Default networks, and the top-20 network breakdown (Figure 5(ii)) confirms the largest contribution comes from the limbic and somatomotor systems. The hemisphere summary (Figure 5(iii)) indicates a modest left-hemisphere predominance (12 left vs 8 right among the top 20), a pattern that may reflect sample composition or lateralized disease but requires subject-level lateralization metadata to interpret definitively. Gradient-based importances (summarized in Figure 5(v)) are relatively tightly distributed — several parcels consistently show higher attribution — while the effect-size versus −log10(p) scatter (Figure 5(iv)) shows that univariate differences are generally small-to-moderate; together these panels imply that the transformer is exploiting subtle, distributed spatiotemporal signals (captured by gradient attributions) that are not always mirrored by large univariate mean shifts.

**Figure 5.**
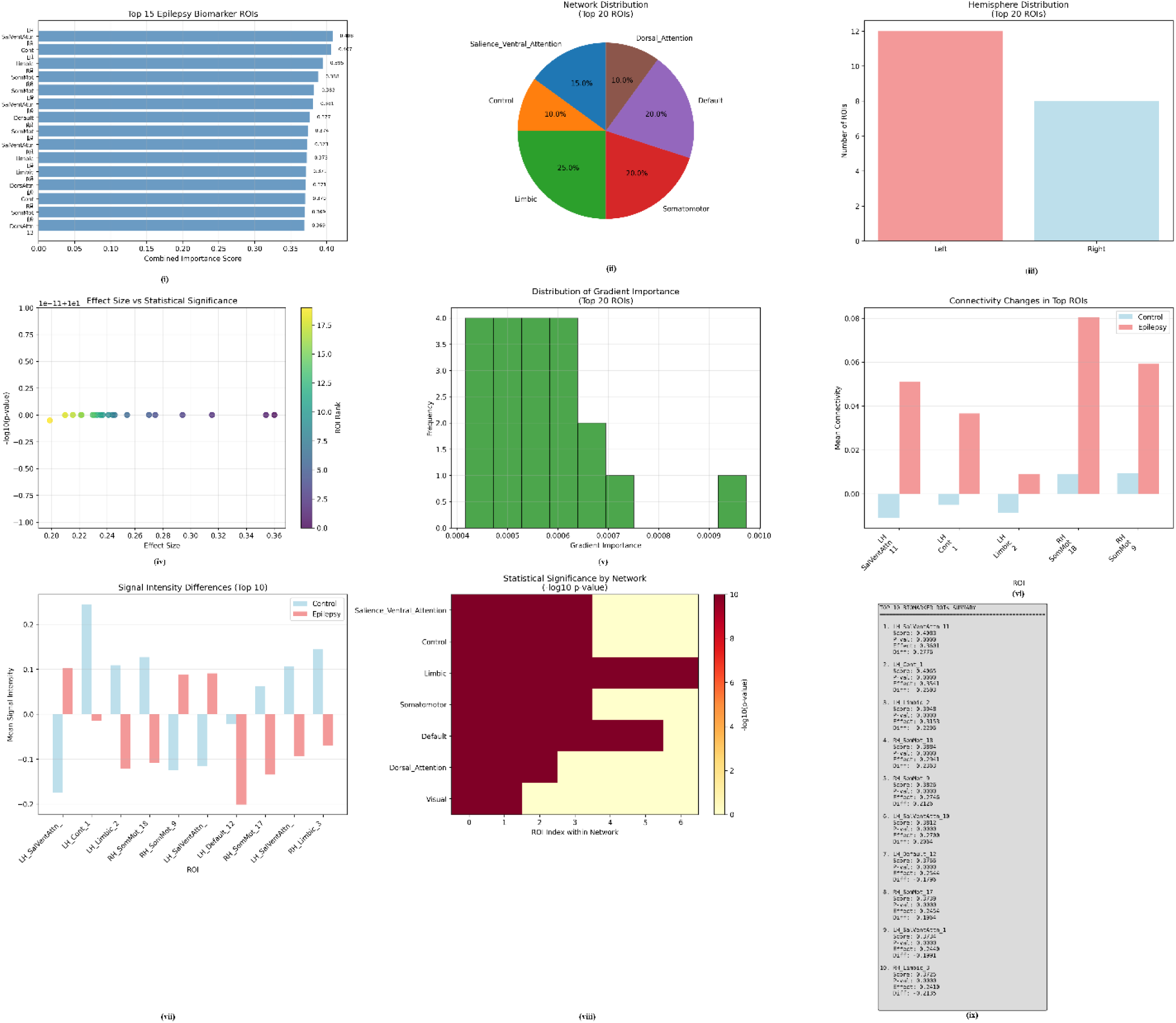
Biomarker discovery from gradient-attribution and statistical analysis (Schaefer-200). (i) Top 15 parcels by combined score (gradient importance + significance + effect size). (ii) Network composition of top 20 ROIs. (iii) Hemisphere counts (top 20). (iv) Effect size vs −log (p) for top parcels. (v) Histogram of gradient-derived importance (top 20). (vi) Mean connectivity (Control vs Epilepsy) for representative top ROIs. (vii) Mean signal differences (top 10 ROIs). (viii) Network-level significance (−log p). (ix) Top-10 ROI summary table (score, p, effect, mean diff).

The network-context analyses and summary table add biological plausibility but also important caveats. Group-average connectivity contrasts (Figure 5(vi)) show that several top parcels (for example LH_SalVentAttn_11) have increased mean coupling in patients relative to controls, and the mean-signal bar chart for top ROIs (Figure 5(vii)) reveals heterogeneous amplitude differences across parcels; the network-level significance heatmap (Figure 5(viii)) and the textual top-10 summary (Figure 5(ix)) provide the numeric details used to prioritize follow-up. Critically, these outputs should be viewed as candidate biomarkers: the attribution scores are model-dependent, the reported p-values in this pipeline come from concatenated timepoints and are not corrected for multiple comparisons, and connectivity estimates depend on preprocessing/denoising choices. For robust inference we therefore recommend reporting FDR-corrected p-values, performing subject-level statistics or permutation/cluster tests, assessing selection stability via bootstrap resampling of the entire attribution + selection pipeline, and seeking external replication and clinical correlation (e.g., with seizure lateralization, EEG, or surgical outcome) before claiming diagnostic validity.

### 3.5 Model Comparison – Proposed Transformer vs Baselines

To contextualize the performance of the proposed regularized transformer, we compared it (mean values reported across 4-fold stratified group cross-validation) to seven alternative deep learning architectures: Multi-Layer Perceptron (MLP), 1D Convolutional Neural Network (CNN-1D), LSTM, hybrid CNN–LSTM, Graph Convolutional Network (GCN), Graph Attention Network (GAT), and an Attention-Only model. Comparative summaries are presented in Table 3.

**Table 3.**
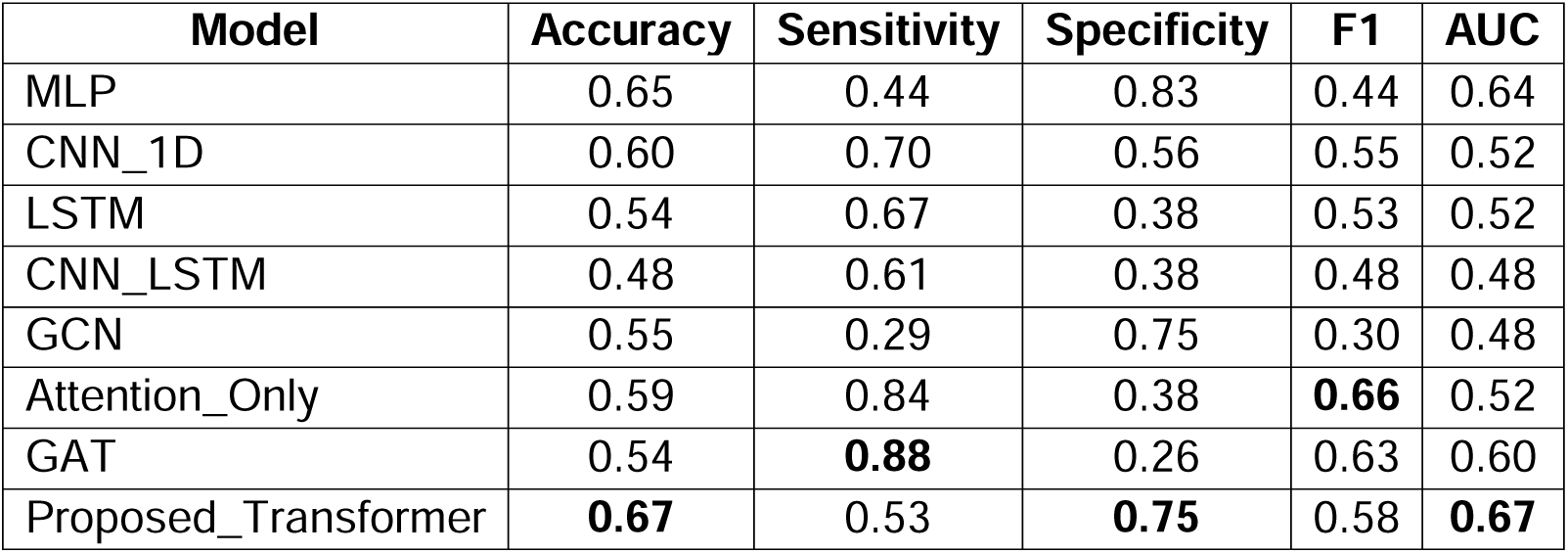
Cross-validated comparison of model performance (mean across 4-fold StratifiedGroupKFold): Accuracy, Sensitivity (recall for epilepsy), Specificity (recall for controls), F1-score and AUC.

Among baselines, the MLP achieved a reasonably balanced profile (accuracy = 0.65, AUC = 0.64, specificity = 0.83) but exhibited limited sensitivity and F1 (both 0.44), indicating it tends to miss positive cases. Temporal architectures (CNN-1D, LSTM) achieved higher sensitivity (0.70 and 0.67, respectively) but at the cost of lower specificity and modest AUCs (≈0.52), suggesting unstable ranking performance across held-out folds. The CNN–LSTM hybrid underperformed on most metrics (accuracy = 0.48, AUC = 0.48), implying that naively stacking spatial and temporal modules did not improve generalization in this dataset. Graph methods showed extreme trade-offs: GAT and the Attention-Only model reached very high sensitivity (0.88 and 0.84) but suffered severe specificity losses (0.26 and 0.38), producing many false positives; the GCN showed the opposite tendency (sensitivity 0.29, specificity 0.75). These asymmetric error profiles are undesirable for a clinically oriented diagnostic tool that must balance detecting cases and avoiding excessive false alarms.

By contrast, the proposed regularized transformer delivered the most balanced performance: accuracy 0.67, AUC 0.67, F1 0.58, sensitivity 0.53 and specificity 0.75. The transformer’s higher AUC and F1 indicate superior ranking ability and a better precision–recall trade-off compared with the baselines, while its specificity remains high enough to limit false alarms. Taken together, these empirical advantages—consistent improvements across multiple metrics—motivated selection of the transformer as the primary model for downstream attribution, biomarker analyses and external testing.

### 3.6 Discussions

In this study, we developed and evaluated a regularized transformer model for classifying epilepsy patients from healthy controls using resting-state fMRI data parcellated with the Schaefer-200 atlas. Our core finding is that the proposed architecture achieved the best-balanced performance on a rigorous 4-fold stratified group cross-validation, yielding a mean AUC and accuracy of 0.67. Notably, the best-performing fold demonstrated the model’s considerable potential, achieving an accuracy of 0.77, an AUC of 0.76, and a particularly strong balance between sensitivity (0.83) and specificity (0.88). This peak performance indicates that with a sufficiently representative and homogeneous training cohort, the model can achieve a clinically promising level of discrimination, effectively identifying both patients and controls with high reliability.

This best-fold performance profile contrasts with the more conservative average cross-validation results (mean sensitivity 0.53, specificity 0.75) and highlights the impact of data variability and limited sample size on model stability. The high performance in one fold suggests that the architecture itself is capable of learning robust features for classification, but the inconsistency across folds underscores the challenge of generalizing from a small dataset where the partitioning of data can significantly influence outcomes.

The model’s performance on an external validation cohort from a different institution underscored a critical challenge in clinical neuroimaging AI: generalizability. While specificity remained high (0.80), indicating a retained ability to correctly identify healthy controls, sensitivity dropped to 0.40. This decline highlights the model’s limited ability to generalize its detection of patient-specific features across different scanners and acquisition protocols. The disparity between the high best-fold results and the external validation performance emphasizes that while the model has high capacity, its practical utility is currently constrained by dataset size and heterogeneity, necessitating domain adaptation techniques and large-scale, multi-site validation studies before any consideration of clinical deployment.

Our use of gradient-based attribution identified a distributed set of brain regions— concentrated in limbic, somatomotor, default-mode, and salience/ventral-attention networks—as driving the model’s decisions. The left-hemispheric predominance of these features is biologically plausible for focal epilepsy. However, these results must be interpreted with caution. Model attributions indicate what features were discriminative for the classifier and are not direct, validated evidence of pathophysiology. The presented biomarkers are exploratory and require rigorous subject-level inferential testing, correction for multiple comparisons (e.g., FDR), and external replication to assert their clinical validity.

Several methodological choices were pivotal to the model’s function. The use of the Schaefer-200 atlas provided an optimal balance of dimensionality reduction and anatomical interpretability. The architecture itself, with its node-embedding block, learned positional scaling, and masked attention mechanisms, effectively learned spatiotemporal relationships in the data. Crucially, the extensive multi-level regularization strategy (including tailored data augmentation, dropout, and weight decay) was essential to mitigate overfitting on our modestly sized clinical cohort, providing a pragmatic framework for pilot-scale studies.

#### Clinical Limitations

Several important clinical limitations must be acknowledged. First, the sample size, while typical for a pilot study, limits the statistical power and stability of the estimates, as evidenced by the variance between the best fold and the cross-validation average. Second, the performance drop on external validation clearly demonstrates that scanner and protocol differences present a significant barrier to real-world application, a problem not addressed in this work. Third, the model was trained on a binary classification task (epilepsy vs. control) and does not account for epilepsy subtype, syndrome, or laterality, which are critical for clinical decision-making. Finally, the biomarker discovery pipeline, while generating plausible hypotheses, is not yet sufficient for clinical inference without further statistical validation.

## 4. Conclusion and Future Work

In conclusion, this study presents a regularized transformer architecture that demonstrates a clinically promising capacity for classifying epilepsy from rs-fMRI data, as evidenced by the strong performance (Acc: 0.77, AUC: 0.76) of its best fold. The model provides a robust, interpretable framework that effectively balances sensitivity and specificity on a pilot-scale cohort and identifies biologically plausible neural network biomarkers. However, the discrepancy between the best-fold results and the average cross-validation performance, coupled with a significant drop in sensitivity during external validation, underscores that the current model remains a proof-of-concept. Its translation to clinical practice is ultimately constrained by dataset size and heterogeneity, highlighting a central challenge in medical AI.

To advance this work, future efforts must prioritize large-scale, multi-site validation to rigorously assess generalizability and enable model calibration. Subsequent research should also focus on integrating domain adaptation techniques to mitigate scanner effects, conducting systematic ablations to quantify each architectural component’s contribution, and implementing rigorous statistical corrections to confirm the validity of the identified biomarkers. If these steps are achieved, this framework serves as a foundational step towards a human-in-the-loop clinical tool that could aid in the objective and interpretable diagnosis of epilepsy.

## Funding Statement

The author(s) declare that financial support was received for the research, authorship, and/or publication of this article. NINDS R01NS125897: Following the BOLD Lightening at Rest Strikes the Seizure Onset Zone!

## Ethics declarations

This study was approved by the Institutional Review Board of the University of North Carolina at Chapel Hill (IRB # 24-0833). All rs-fMRI datasets were obtained under informed consent from participants, and all methods were carried out in accordance with relevant guidelines and regulations.

## Data Availability

All data produced in the present study are available upon reasonable request to the authors

## Notes

### Competing Interest Statement

The authors have declared no competing interest.

